# Objective Response by mRECIST is an Independent Prognostic Factor for Overall Survival in Intermediate Hepatocellular Carcinoma Undergoing Transarterial Chemoembolization

**DOI:** 10.1101/2025.10.05.25337382

**Authors:** Xiaoli Jia, Wenjun Wang

## Abstract

**Background:** The prognostic value of objective response assessed by the modified Response Evaluation Criteria in Solid Tumors (mRECIST) remains insufficiently characterized in patients with intermediate-stage hepatocellular carcinoma (HCC) patients undergoing transarterial chemoembolization (TACE).

**Aim:** This study aimed to evaluate the association between objective response and overall survival in this patient population.

**Methods:** A retrospective analysis was conducted on 812 patients with Barcelona Clinic Liver Cancer (BCLC) stage B HCC who received TACE as first-line therapy at a tertiary hospital in China between January 2007 and May 2012. Patients were classified as responders (complete or partial response) or non-responders (stable or progressive disease) based on mRECIST criteria at the first follow-up after TACE. Baseline characteristics, including tumor size, number, and alpha-fetoprotein levels, were recorded. Overall survival was estimated using Kaplan-Meier analysis, and potential prognostic factors were evaluated using Cox proportional hazards models.

**Results:** The objective response rate according to mRECIST was 63.2% (513/812). Responders demonstrated significantly longer median overall survival compared to non-responders (44.0 months vs. 10.9 months; hazards ratio [HR]: 0.31, 95% CI: 0.26–0.38; p < 0.0001). Landmark analysis confirmed the survival advantage among responders (HR: 0.31, 95% CI: 0.26–0.37; p < 0.0001). Multivariable Cox regression analysis identified objective response as an independent prognostic factor for overall survival (HR: 0.37, 95% CI: 0.24–0.57; p < 0.0001), along with tumor size (HR: 1.94, 95% CI: 1.54–2.43; p < 0.0001), tumor number (HR: 1.39, 95% CI: 1.14-1.70; p = 0.001), and alpha-fetoprotein level (HR: 1.23, 95% CI: 1.01–1.49; p = 0.04).

**Conclusion:** Objective response assessed by mRECIST is a robust independent prognostic factor for overall survival in intermediate-stage HCC patients treated with TACE. This finding highlights the clinical importance of achieving an early objective response in improve long-term outcomes. Further prospective studies are needed to validate this result and explore strategies to enhance response rates to TACE.

## Introduction

Hepatocellular carcinoma (HCC) is the most common primary malignancy of the liver and ranks as the sixth most frequently diagnosed cancer worldwide, contributing substantially to cancer-related mortality.^1^ The Barcelona Clinic Liver Cancer (BCLC) staging system is widely adopted to guide therapeutic decision-making in HCC.^2^ Patients with BCLC stage B disease (intermediate-stage HCC) are typically characterized by the presence of multinodular tumors without macrovascular invasion or extrahepatic metastasis. Transarterial chemoembolization (TACE) is currently the standard of care for this patient population, aiming to induce tumor necrosis through selective delivery of chemotherapeutic agents combined with embolic materials to the tumor vasculature.^3^ Despite its widespread use, clinical outcomes following TACE remain heterogeneous,^4^ and the identification of reliable prognostic markers is essential for optimizing patient stratification and treatment planning.

Objective response, assessed using the modified Response Evaluation Criteria in Solid Tumors (mRECIST), has emerged as a potential predictor of overall survival in patients with HCC.^5, 6^ The mRECIST criteria, specifically developed for HCC, evaluate tumor response based on viable tumor burden, thereby offering a more accurate assessment of treatment efficacy compared to the traditional RECIST criteria.^5^ Previous studies have indicated that achieving a complete response or partial response following TACE is associated with improved survival outcomes.^7^ However, the prognostic value of objective response by mRECIST in a large, real-world cohort of patients with BCLC stage B HCC treated with TACE remains insufficiently investigated.

This retrospective study aimed to evaluate the association between objective response assessed by mRECIST and overall survival in patients with intermediate-stage HCC undergoing TACE as first-line therapy. By analyzing a large cohort from a tertiary cancer center, we aimed to determine whether objective response serves as an independent prognostic factor for survival and to identify additional baseline characteristics that may influence outcomes. The findings of this study may provide important insights into the clinical utility of mRECIST in guiding treatment decisions and predicting prognosis in this patient population.

## Material and methods

### Study Design and Patient Selection

This retrospective cohort study analyzed patients diagnosed with intermediate-stage (BCLC stage B) HCC who underwent TACE as first-line treatment at Sun Yat-sen University Cancer Center between January 2007 and May 2012.^8^ The initial screening included 5,005 consecutive HCC cases, from which we selected those meeting the following criteria: confirmed BCLC stage B HCC without evidence of vascular invasion or extrahepatic metastasis, availability of complete baseline clinical and imaging data, no prior history of other malignancies, and TACE as the initial treatment. After excluding patients with insufficient follow-up for response assessment, the final cohort included 812 individuals. The sample selection flow is shown in Figure 1.

**Figure 1.**
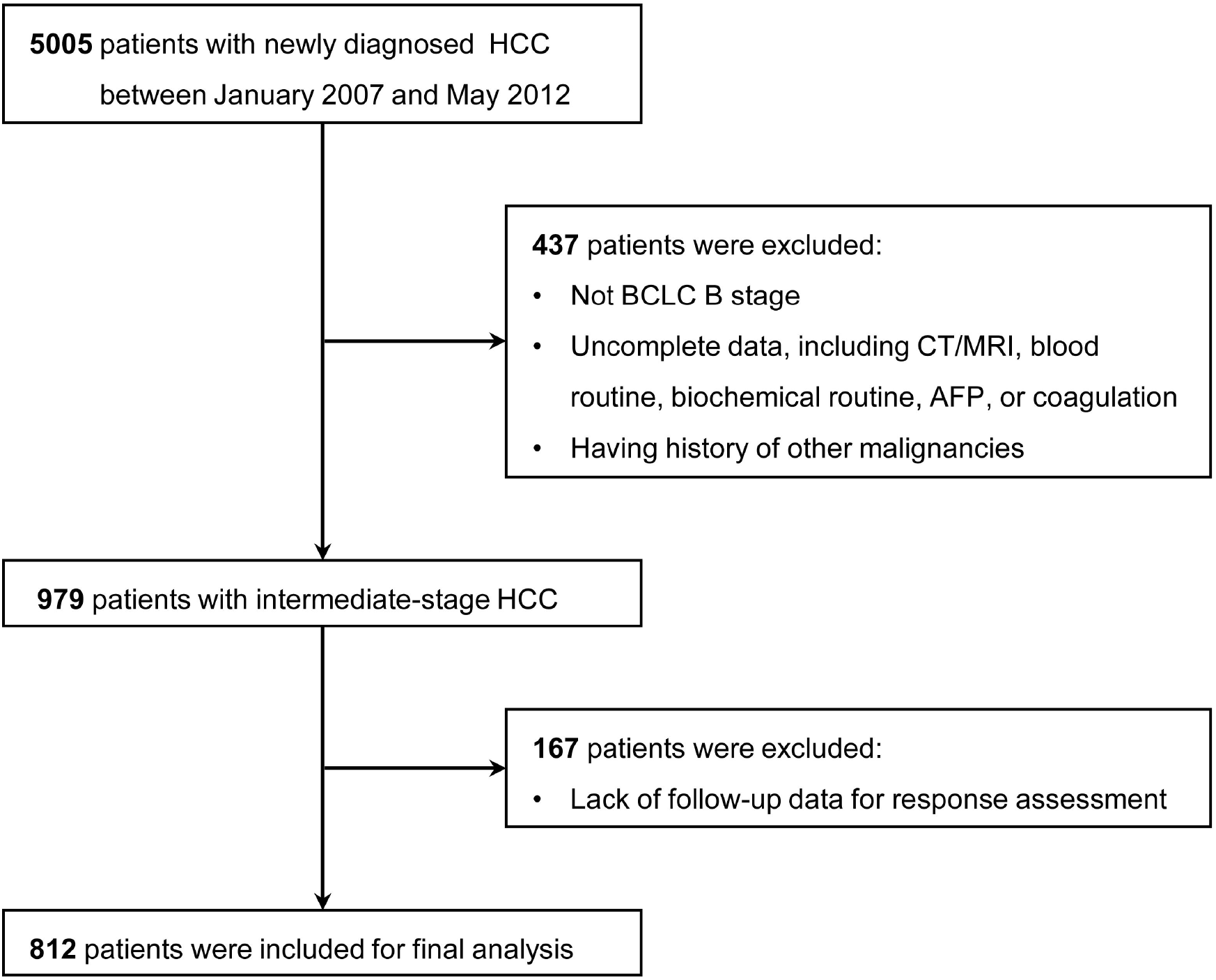
Determination of the study sample.

### Data Collection and Clinical Variables

Baseline demographic and clinical characteristics were systematically extracted from electronic medical records. Key variables included age, sex, etiology of underlying liver disease (hepatitis B or C virus infection), liver function parameters (serum albumin, serum bilirubin, Child-Pugh class, and ALBI grade), tumor characteristics (maximum diameter and number of lesions), and serum alpha-fetoprotein (AFP) levels. Imaging studies, including contrast-enhanced CT or MRI, were reviewed to assess tumor burden and response to treatment.

### Data Collection and Clinical Variables

Baseline demographic and clinical characteristics were systematically extracted from electronic medical records. Key variables included age, sex, etiology of underlying liver disease (hepatitis B or C virus infection), liver function parameters (serum albumin, serum bilirubin, Child-Pugh class, and ALBI grade), tumor characteristics (maximum diameter and number of lesions), and serum alpha-fetoprotein (AFP) levels. Imaging studies, including contrast-enhanced CT or MRI, were reviewed to assess tumor burden and response to treatment.

### Treatment and Follow-up

All patients received TACE according to institutional protocols, which involved the administration of chemotherapeutic agents (doxorubicin or cisplatin) mixed with lipiodol or drug-eluting beads, followed by embolization of the tumor-feeding arteries. Treatment sessions were repeated on demand based on tumor response and liver function.

Post-treatment follow-up followed a standardized schedule, with clinical and imaging evaluations conducted monthly during the initial treatment phase. For patients achieving complete remission, follow-up visits were scheduled every 2–3 months for the first two years and every 3–6 months thereafter. Imaging was conducted using dynamic contrast-enhanced CT or MRI and was assessed by investigators to evaluate treatment response according to mRECIST criteria.

### Outcome Measures and Response Assessment

The primary endpoint was overall survival, defined as the time interval from the first TACE procedure to death from any cause. Tumor response was evaluated at the first follow-up imaging examination, typically 6–8 weeks after TACE, and categorized according to mRECIST criteria. Patients were classified as responders if they achieved a complete response or partial response, while those with stable disease or progressive disease were classified as non-responders.

### Statistical Analysis

Statistical analyses were performed using STATA version 15.0. Categorical variables were compared between responders and non-responders using Fisher’s exact test, while continuous variables were analyzed with the Mann-Whitney U test. Survival probabilities were estimated using the Kaplan-Meier method, and differences between groups were assessed with the Mantel-Byar test to account for the time-dependent nature of treatment response.

To minimize guarantee-time bias, a landmark analysis was conducted at the time of the first follow-up assessment. Additionally, Cox proportional hazards regression models were employed to identify independent predictors of survival. Univariable analysis was performed for all baseline variables, and those with a p-value less than 0.10 were included in the multivariable model. Objective response was treated as a time-dependent covariate in the Cox regression to accurately capture its dynamic impact on survival.

The study was approved by the Institutional Review Board of Second Affiliated Hospital of Xi’an Jiaotong University (2025-001). This study is a secondary analysis of the original data from Sun Yat-sen University Cancer Center (approval number: 2017-FXY-129).^8^ Given the retrospective nature of the study, the requirement for informed consent was waived. Patient data were anonymized to ensure confidentiality.

## Results

At the end of follow-up, 440 of the 812 patients treated with TACE had died, with a median OS of 27.2 months (95% CI, 23.9–32.0). At the first follow-up after TACE, 513 (63.2%) were responders and 299 (36.8%) were non-responders. Baseline characteristics of responders and non-responders are shown in Table 1.

**Table 1.**
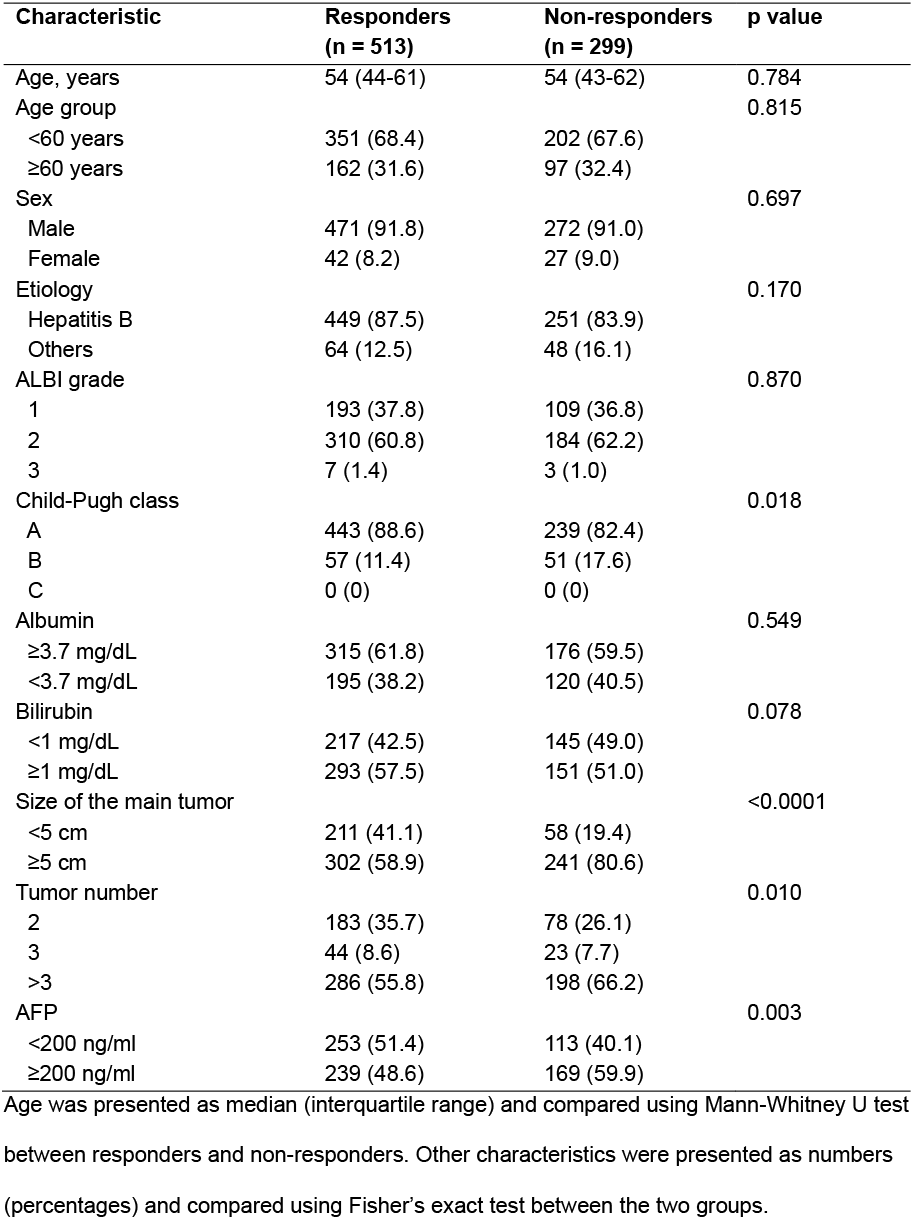
Baseline characteristics of responders and non-responders.

| Characteristic | Responders<br>(n = 513) | Non-responders<br>(n = 299) | p value |
| --- | --- | --- | --- |
| Age, years | 54 (44-61) | 54 (43-62) | 0.784 |
| Age group |  |  | 0.815 |
| <60 years | 351 (68.4) | 202 (67.6) |  |
| ≥60 years | 162 (31.6) | 97 (32.4) |  |
| Sex |  |  | 0.697 |
| Male | 471 (91.8) | 272 (91.0) |  |
| Female | 42 (8.2) | 27 (9.0) |  |
| Etiology |  |  | 0.170 |
| Hepatitis B | 449 (87.5) | 251 (83.9) |  |
| Others | 64 (12.5) | 48 (16.1) |  |
| ALBI grade |  |  | 0.870 |
| 1 | 193 (37.8) | 109 (36.8) |  |
| 2 | 310 (60.8) | 184 (62.2) |  |
| 3 | 7 (1.4) | 3 (1.0) |  |
| Child-Pugh class |  |  | 0.018 |
| A | 443 (88.6) | 239 (82.4) |  |
| B | 57 (11.4) | 51 (17.6) |  |
| C | 0 (0) | 0 (0) |  |
| Albumin |  |  | 0.549 |
| ≥3.7 mg/dL | 315 (61.8) | 176 (59.5) |  |
| <3.7 mg/dL | 195 (38.2) | 120 (40.5) |  |
| Bilirubin |  |  | 0.078 |
| <1 mg/dL | 217 (42.5) | 145 (49.0) |  |
| ≥1 mg/dL | 293 (57.5) | 151 (51.0) |  |
| Size of the main tumor |  |  | <0.0001 |
| <5 cm | 211 (41.1) | 58 (19.4) |  |
| ≥5 cm | 302 (58.9) | 241 (80.6) |  |
| Tumor number |  |  | 0.010 |
| 2 | 183 (35.7) | 78 (26.1) |  |
| 3 | 44 (8.6) | 23 (7.7) |  |
| >3 | 286 (55.8) | 198 (66.2) |  |
| AFP |  |  | 0.003 |
| <200 ng/ml | 253 (51.4) | 113 (40.1) |  |
| ≥200 ng/ml | 239 (48.6) | 169 (59.9) |  |

Age was presented as median (interquartile range) and compared using Mann-Whitney U test between responders and non-responders. Other characteristics were presented as numbers (percentages) and compared using Fisher's exact test between the two groups.

Objective response rate assessed by mRECIST was 63.2% (513/812) in the study population. The median overall survival was 44.0 months (95% CI, 38.7–54.1) in responders and 10.9 months (9.8-14.4) in non-responders. The HR was 0.31 (95% CI, 0.26–0.38; P <0.0001). Kaplan-Meier estimates for responders and non-responders in the study population are shown in Figure 2.

**Figure 2.**
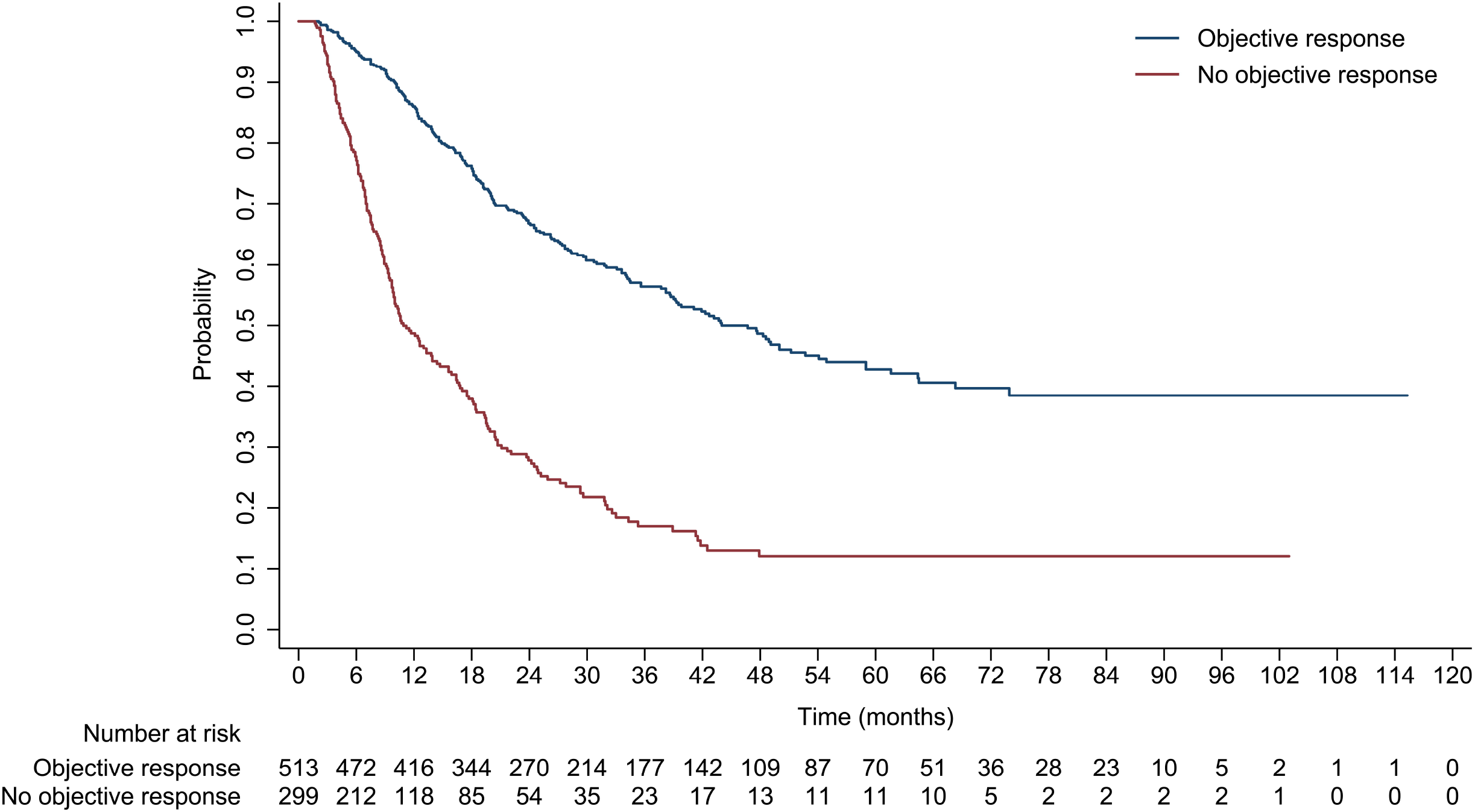
Overall survival by objective response assessed by mRECIST in intermediate-stage HCC patients treated with TACE.

Landmark analyses at the time of the first follow-up after TACE also showed that overall survival was significantly longer in responders than in non-responders (HR, 0.31, 95% CI, 0.26– 0.37; p <0.0001) (Figure 3).

**Figure 3.**
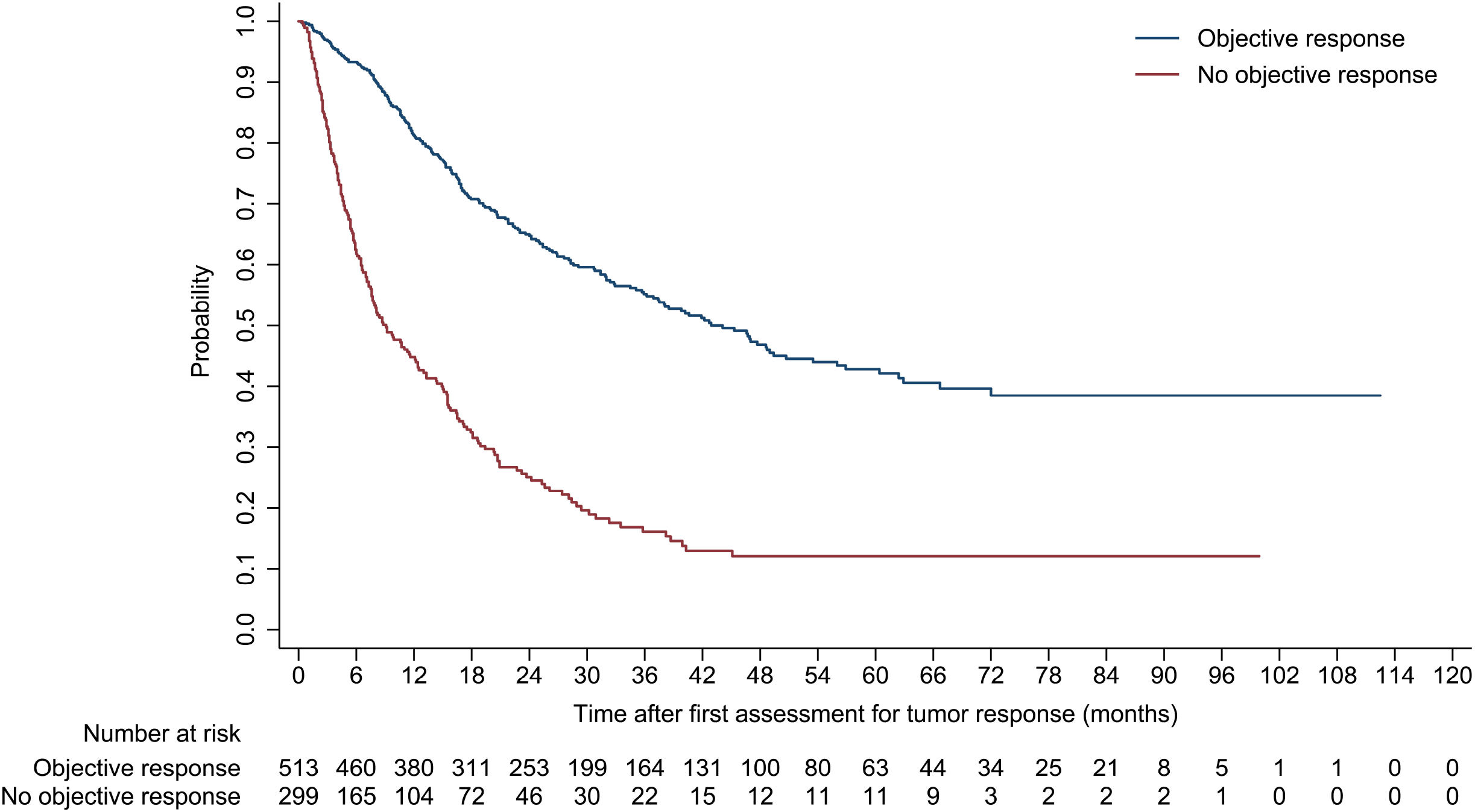
Landmark analyses for overall survival by objective response assessed by mRECIST in intermediate-stage HCC patients treated with TACE.

Multivariable Cox regression analysis revealed that objective response assessed by mRECIST was an independent prognostic factor (HR, 0.37, 95% CI, 0.24–0.57; p < 0.0001). Other prognostic factors identified were tumor size (HR, 1.94, 95% CI, 1.54–2.43; p < 0.0001), tumor number (HR, 1.39, 95% CI, 1.14–1.70; p = 0.001), and AFP level (HR, 1.23, 95% CI, 1.01– 1.49; p = 0.04] (Table 2).

**Table 2.**
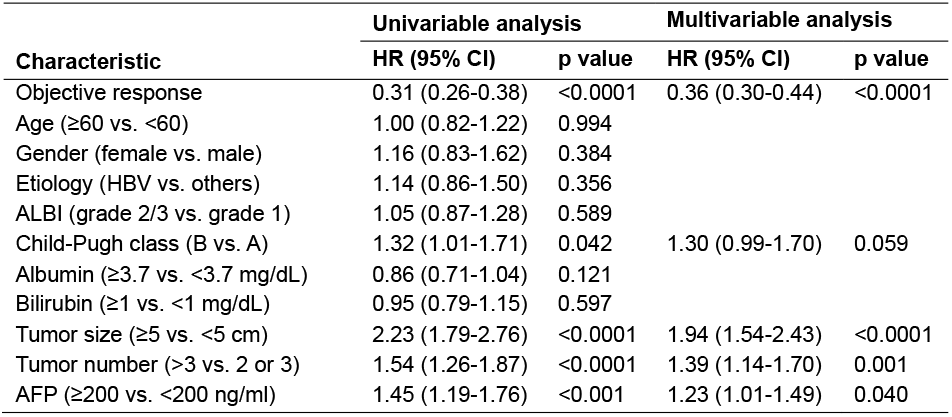
Factors associated with OS in intermediate HCC treated with TACE.

## Discussion

This retrospective analysis, conducted within a large tertiary care center, demonstrated a significant association between investigator-assessed objective response by mRECIST and overall survival in HCC patients treated with TACE. The results confirm that objective response, as defined by mRECIST, serves as an independent prognostic factor in this clinical setting. Patients who achieved a complete or partial response exhibited markedly improved survival compared to those with stable or progressive disease.

To our knowledge, this study represents the largest cohort to date, encompassing 812 patients, evaluating the predictive validity of mRECIST for survival outcomes in TACE-treated HCC. Previous studies, though consistent in direction, were limited by smaller sample sizes (ranging from 83 to 368 patients) and did not adequately account for guarantee-time bias.^7^ Although reported hazard ratios in earlier studies were broadly consistent with our findings (ranging from 0.15 to 0.71 for responders versus non-responders), the methodological approach presented here provides a more rigorous adjustment for time-related biases.

To mitigate guarantee-time bias, the analysis employed two well-established statistical approaches: landmark analysis at the first post-TACE follow-up and a multivariable Cox regression model treating objective response as a time-dependent covariate.^9-11^ These methodologies effectively minimize bias arising from temporal variations in response evaluation.^9-11^ The observation that response status assessed early during treatment serves as a robust prognostic indicator highlights the critical impact of initial tumor response on long-term clinical outcomes.

In addition to objective response, the analysis confirmed several previously recognized prognostic factors, including tumor size, tumor number, and baseline AFP levels, further validating the multidimensional nature of outcome prediction in this patient population.^4^

Several limitations should be acknowledged. First, the lack of detailed treatment data, such as specific TACE techniques (conventional vs. drug-eluting beads), times of TACE, combination therapies, and therapies after progression, restricts more depth interpretation. The most recent evidence supports combination of TACE with systemic therapy for unresectable HCC, especially for those with heavy tumor burden.^12-15^ Future studies should evaluate the predictive value in this setting. Second, the single-center retrospective design may limit the generalizability of the findings. Multi-institutional and longitudinal studies would strengthen these conclusions.

## Conclusion

In summary, this study reinforces the value of mRECIST as an early and reliable indicator of survival in intermediate-stage HCC patients undergoing TACE. These findings support the integration of standardized response assessment into clinical decision-making and highlight the need for response-adapted treatment strategies. Further validation through prospective, multi-center studies and in the setting of combination with systemic therapy will help consolidate the role of mRECIST as a cornerstone of outcome prediction in HCC management.

## Data Availability

The dataset used in the study can be freely obtained from the Dryad Digital Repository: https://datadryad.org/dataset/doi:10.5061/dryad.pd44k8r

## Data Sharing Statement

The dataset (doi:10.5061/dryad.pd44k8r) can be freely obtained from the Dryad Digital Repository (https://datadryad.org).

## Ethics Approval and Informed Consent

The study was approved by the Institutional Review Board of Second Affiliated Hospital of Xi’an Jiaotong University (2025-001). This study is a secondary analysis of the original data from Sun Yat-sen University Cancer Center (approval number: 2017-FXY-129). Given the retrospective nature of the study, the requirement for informed consent was waived. Patient data were anonymized to ensure confidentiality.

## Author Contributions

Wang and Jia take responsibility for the integrity of the data and the accuracy of the data analysis. Concept and design: Wang, Jia. Analysis and interpretation of data: Wang, Jia. Drafting of the manuscript: Wang, Jia. Critical revision of the manuscript for important intellectual content: Wang, Jia.

## Disclosure

The authors report no conflicts of interest in this work.

## Notes

### Competing Interest Statement

The authors have declared no competing interest.

### Funding Statement

This study did not receive any funding.

### Author Declarations

The study was approved by the Institutional Review Board of Second Affiliated Hospital of Xi'an Jiaotong University (2025-001). This study is a secondary analysis of the original data from Sun Yat-sen University Cancer Center (approval number: 2017-FXY-129).

